# Longitudinal modelling of clonal hematopoiesis reveals altered early clonal dynamics in people with HIV

**DOI:** 10.64898/2026.04.08.26350407

**Authors:** Valeriia Timonina, Alexandra Calmy, Matthias Cavassini, Gioele Capoferri, Huldrych F. Günthard, Laura N. Walti, Patrick Schmid, Philip E. Tarr, Alexander G. Bick, Jacques Fellay, the Swiss HIV Cohort Study (SHCS)

## Abstract

Clonal hematopoiesis of indeterminate potential (CHIP) is an age-associated condition linked to chronic inflammation, cardiovascular diseases, and hematological malignancies. People with HIV (PWH) show higher CHIP prevalence than the general population, but whether this is due to earlier emergence, altered clonal expansion dynamics, or differences in selective pressures is unclear.

We reconstructed longitudinal CHIP trajectories in 52 PWH using serial blood samples spanning up to 25 years and identified patterns of CHIP dynamics consistent with known gene-specific fitness patterns. Larger clone size was associated with lower CD4 T-cell count and lower CD4/CD8 ratio. Compared with a general population cohort, PWH showed higher VAF across the observed age range and steeper early trajectory increases, while long-term expansion rates were broadly similar. These findings support a model in which HIV-associated immune dysregulation alters the hematopoietic fitness landscape, contributing to earlier clonal expansion and increased burden of CHIP in PWH.

## Introduction

Clonal Hematopoiesis of Indeterminate Potential (CHIP) refers to the expansion of hematopoietic stem and progenitor cell clones carrying leukemia-associated somatic mutations at variant allele frequency (VAF) ≥2% in individuals without overt hematological malignancy. CHIP prevalence increases markedly with age, reaching 20-40% in individuals older than 70 years. CHIP promotes a pro-inflammatory phenotype in hematopoietic and innate immune cells, creating a positive feedback loop between inflammation and clonal expansion that contributes to hematopoietic and systemic aging^1,2^. Consequently, CHIP has been associated with an increased risk of cardiovascular disease^3,4^, chronic kidney and liver disease^5,6^, and osteoporosis^7,8^, supporting a broader role for mutant hematopoietic clones in age-related morbidity^9,10^.

To reconstruct temporal trajectories of CHIP clones and model their growth dynamics, several recent studies have moved beyond cross-sectional epidemiological analyses to track clonal hematopoiesis within individuals longitudinally^11–14^. By measuring the variant allele frequency (VAF), a proxy for clone size, across serial blood samples, it is possible to reconstruct temporal trajectories of CHIP clones and model their growth dynamics. These longitudinal approaches enable estimation of clonal fitness and provide insight into the biological behavior and potential clinical relevance of specific CHIP variants. For example, Fabre et al.^11^ demonstrated that CHIP clones grow at gene-specific rates, with mutations in *TET2*, *ASXL1*, and *PPM1D* generally expanding faster than those in *DNMT3A* and *TP53*, and that clonal fitness effects are largely consistent across individuals, suggesting intrinsic differences in mutation-driven selective advantage. These CHIP variant-defined fitness showed consistency across different cohorts^13^.

People with HIV (PWH) exhibit features of accelerated biological aging, including earlier onset of age-related comorbidities^15,16^, despite effective antiretroviral therapy (ART) and sustained viral suppression^17^. Several studies have reported an increased prevalence of CHIP in PWH compared with the general population^18,19^. Proposed explanations include increased background inflammation, exposure to anti-HIV treatment, and immune dysregulation^18–22^. Yet, it remains unclear whether increased CHIP prevalence reflects earlier acquisition of CHIP driver mutations, faster expansion of clones, differences in selective pressures acting on hematopoietic stem cells, or a combination of these factors.

Biological alterations associated with HIV infection may influence both the emergence and expansion of CHIP clones. Persistent immune activation, altered cytokine signalling, and impaired immune surveillance may give a selective advantage to mutant hematopoietic stem cells. This could lead to earlier detectable clonal emergence, higher VAF at similar ages, or modified growth dynamics relative to the general population. Conversely, ART immune reconstitution may have compensatory effects. Longitudinal trajectory features such as estimated age of origin and growth rate may therefore provide insight into how HIV-associated immune perturbations shape clonal hematopoiesis.

Here, we reconstruct longitudinal CHIP trajectories spanning up to 25 years in PWH enrolled in the Swiss HIV Cohort Study (SHCS) and compare these trajectories with those observed in the SardiNIA population cohort. We aim to characterize patterns of clonal growth, identify clinical and immunological correlates of clonal expansion, and assess whether CHIP dynamics differ systematically between PWH and the general population.

## Methods

### Participant samples and ethics

Our study population includes PWH enrolled in the SHCS, a multicentre, open-label, longitudinal, prospective cohort study in Switzerland, which has recruited more than 22,000 PWH since 1988. The SHCS was approved by the ethics committees of the participating institutions, and written informed consent was obtained from all participants^23^.

Among SHCS participants aged 60 years or older previously screened for CHIP and identified as CHIP carriers^20^, we selected 54 individuals with available historical Peripheral Blood Mononuclear Cells (PBMC) samples spanning at least 20 years. For each participant, 5–6 serial PBMC samples were selected at approximately five-year intervals before and, when available, after the initial CHIP screening, yielding up to 25 years of longitudinal follow-up.

### CHIP identification

Targeted, high-depth DNA sequencing for CHIP detection was conducted at the Vanderbilt Technologies for Advanced Genomics (VANTAGE) core facility using Twist Biosciences’ hybrid capture platform. This approach enabled the sequencing of 24 genes implicated in CHIP (commonly referred to as “CHIP genes”) at a coverage exceeding 500×, facilitating reliable detection of low-frequency somatic variants^24^. The gene panel includes *ASXL1, ASXL2, BRCC3, CBL, DNMT3A, ETNK1, GNAS, GNB1, IDH1, IDH2, JAK2, KIT, KRAS, MPL, NRAS, PPM1D, SETBP1, SF3B1, SRSF2, TET2, TP53, U2AF1, ZBTB33,* and *ZNF318*. Altogether, these genes—or selected regions within them known to harbor CHIP-associated variants—span approximately 45kb of genomic sequence. Analysis of this panel enables the detection of over 95% of known CHIP variants^24,25^.

Sequencing reads were first aligned to the GRCh38 reference genome using BWA-MEM. CHIP-associated variants were then identified in each individual using GATK MuTect2 in combination with manual curation, following previously described methods^18,25^. Variants were retained if they met the following criteria: a minimum sequencing depth of 20 reads, at least three reads supporting the alternative allele, and evidence from both forward and reverse strands.

### Trajectories dynamics description

CHIP trajectories were defined as time series of VAF measurements for individual variants detected at at least two timepoints, with at least one measurement ≥2% VAF. We assumed a VAF of 0 when no variant was identified in the sequenced sample.

We fitted cubic splines to the measured VAF against age and predicted VAF values at 6-month intervals. First and second derivatives of the fitted curves were used as descriptive measures of clonal growth dynamics (Figure 1B), as follows:

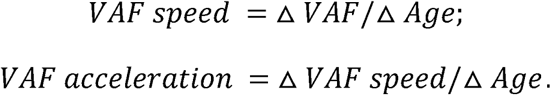

**Figure 1.**
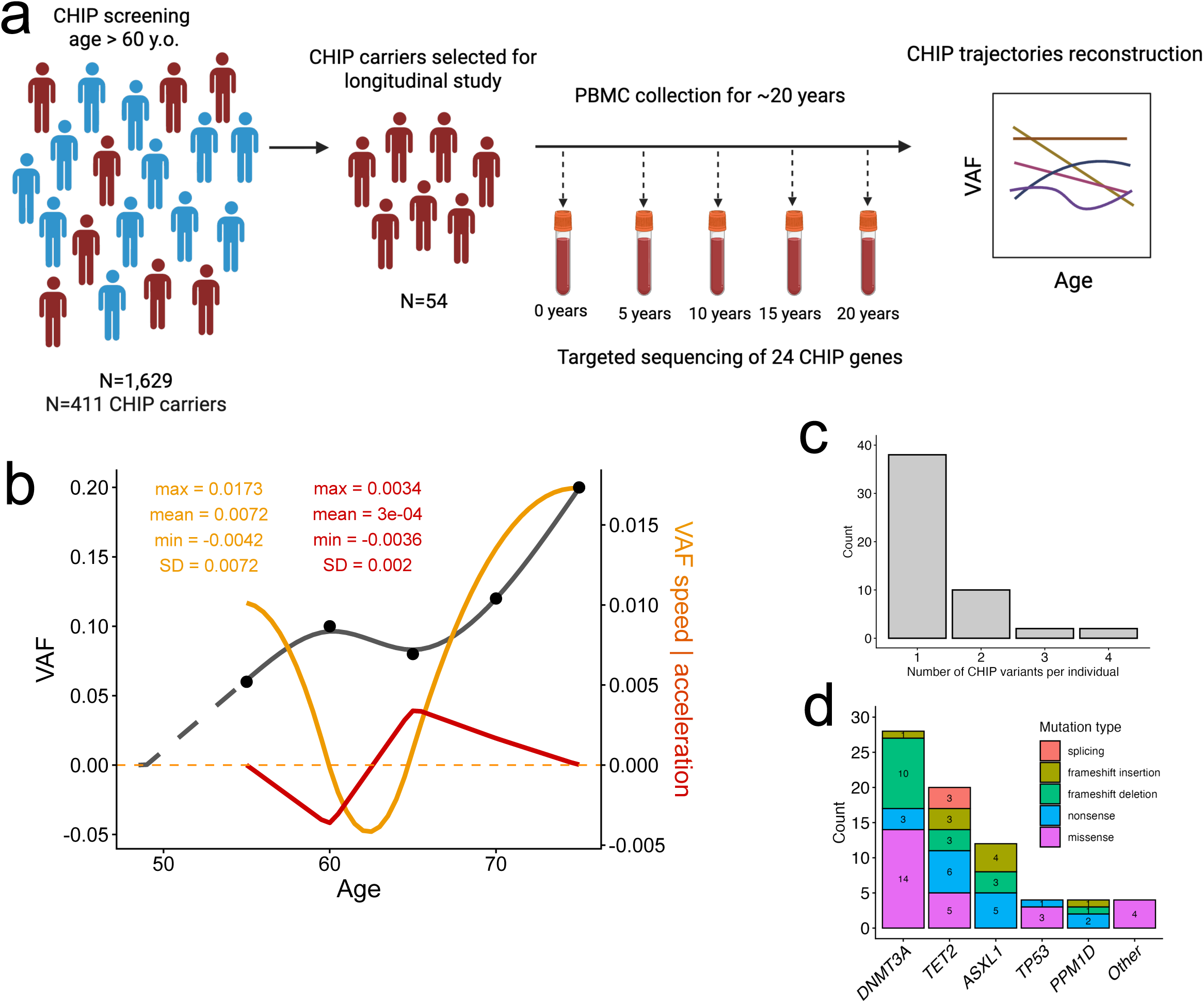
Longitudinal CHIP study design, trajectories, and mutation profiles in PWH. a. The sample selection outline: we selected 54 CHIP carriers from 1,629 SHCS participants >60 years for PBMC-based targeted sequencing of 24 CHIP genes across 5-6 timepoints spanning 20-25 years to reconstruct CHIP VAF trajectories. b. An example of one longitudinal CHIP trajectory reconstruction. We fitted a cubic smoothing spline to the VAF measurements (dark grey), then calculated the longitudinal first (yellow) and second (red) derivatives to estimate VAF speed and acceleration, respectively. For the analyses, we used both longitudinal estimated and derived summary statistics, such as mean, minimum (min), maximum (max), and standard deviation (SD). Age of origin was estimated by linear extrapolation of the VAF trajectory (dashed grey line) when possible and when not already available from measured data. c. Number of CHIP variants per individual. d. CHIP variant distribution by gene and mutation type.

This way, we got longitudinal measures of VAF, VAF speed, and VAF acceleration with values every 6 months. For longitudinal measures, we also derived summary statistics for each trajectory, that is, mean, minimum, maximum, and standard deviation. When possible, we estimated the age of detectable clonal origin, that is, the age when VAF was 0%, by linear interpolation of fitted trajectories.

### Trajectories clustering

Longitudinal CHIP VAF trajectories were aligned and clustered using dynamic time warping (DTW) to account for heterogeneity in sampling time and trajectory length across individuals. VAF was standardized within individuals using z-normalization to ensure comparability of temporal patterns. Clustering was performed with the tsclust() function using the dtw_basic distance and DTW barycenter averaging (centroid = “dba”), with L2 normalization (norm = “L2”) and a Sakoe–Chiba warping window of 20 (window.size = 20) ^26^. The optimal number of clusters was evaluated using internal cluster validity indices. For the number of clusters ranging from 2 to 10, the validity indices were calculated using the cvi() function from dtwclust and compared graphically across cluster numbers. The final clustering solution was selected on the basis of these indices together with visual inspection of cluster separation. Clustering stability was assessed using the leave-one-out method, when performing the clustering of all the trajectories except one and then predicting the cluster for the left-out trajectory.

### Association with longitudinal blood parameters

To assess the longitudinal association between hematologic parameters and CHIP dynamics, we fitted linear mixed-effects models using the lme4 package in R. Separate models were constructed for each combination of CHIP trajectory metric (VAF, VAF speed, and VAF acceleration) and blood parameter. Analyses were restricted to timepoints following ART initiation. Age was standardized (z-score), and blood parameters were log10-transformed. VAF values were additionally log10-transformed to account for skewness. For each outcome–predictor pair, a set of candidate models including fixed effects for age, the blood parameter, and their interaction was fitted with either random intercepts or random intercepts and slopes for age at the individual level. Model selection was guided by the Akaike Information Criterion (AIC) and marginal and conditional R² values calculated using the performance package. Model diagnostics were assessed through inspection of residual distributions and fitted values.

### Comparison with the general population cohort

#### Matching of trajectories

To compare CHIP trajectories of PWH from SHCS to non-infected individuals, we utilized publicly available data from the SardiNIA longitudinal cohort. For all analyses, in both cohorts, we kept only trajectories in the 5 most common genes in SHCS - *DNMT3A*, *TET2*, *ASXL1*, *PPM1D*, and *TP53*. To account for the difference in age between SHCS and SardiNIA, we performed several levels of filtering and matching - 1) completely unmatched; 2) age-filtered to keep only time points intersecting between the two cohorts; 3) 1:1 matching where for each CHIP trajectory in SHCS we selected the trajectory in Sardinia in the same gene that is the closest in terms of age; 4) matched trajectories that were also age-filtered to keep only time points intersecting between the two cohorts (Figure S6). To increase statistical power, we used unmatched trajectories for the main analysis, and the rest were used for the sensitivity analysis to explore the robustness and consistency of the associations.

#### Mixed linear models

To compare longitudinal CHIP trajectories between the SHCS and SardiNIA cohorts, we fitted linear mixed-effects models using the lme4 package in R. We modelled log10(VAF) as a function of age and HIV status (SHCS vs SardiNIA), including either a random intercept and random slope for age at the individual trajectory level. We also fitted a model including the interaction of age and HIV status to test for differences in slopes between the two cohorts. Model fit was assessed by visual inspection of residual and fitted-value plots.

#### Clustering

Trajectories from the SardiNIA cohort were assigned to the clusters derived from SHCS data using DTW. The DTW-based clustering model trained on SHCS trajectories (tsclust, dtwclust package, see above) was applied to SardiNIA trajectories using the predict() function, which assigns each new trajectory to the nearest cluster centroid based on DTW distance. Each SardiNIA trajectory was thus classified according to its similarity to the SHCS-derived cluster profiles.

#### DBA

To compare the average trajectory shape between the SHCS and SardiNIA cohorts, we computed cohort-specific dynamic time warping barycenters using the DBA (DTW barycenter averaging) algorithm implemented in the dtwclust R package. For each cohort, the set of CHIP VAF trajectories was used as input, and the longest trajectory (with maximum follow-up time) from that cohort was provided as the initial centroid. DBA was run for up to 20 iterations to obtain a representative average trajectory for each cohort. The resulting SHCS and SardiNIA barycenters were then plotted and compared over aligned pseudo-time to visualize differences in the overall temporal pattern of CHIP dynamics between cohorts.

## Results

### The landscape of CHIP trajectories in PWH

Of 1,629 SHCS participants aged 60 years or older who had previously been screened for CHIP, 411 were identified as CHIP carriers ^20^. We selected 54 of them to study the longitudinal dynamics of CHIP variants, based on the availability of PBMCs for at least 20 years of follow-up. For each study participant, we sequenced 5 or 6 PBMC samples collected at 5-year intervals (Fig. 1A). The identification of CHIP trajectories was based on the detection of the same CHIP variant at >=2 time points, with at least one of them with VAF >= 2%. This resulted in 72 trajectories with reconstructed longitudinal dynamics (Fig. 1B) in 52 individuals (Fig. S1). The characteristics of the 52 individuals with at least one CHIP trajectory are presented in Table 1.

**Table 1.** Epidemiological description of 52 SHCS participants left after filtering CHIP trajectories.

|  |  |
| --- | --- |
| **min Age, years, median (IQR)** | 54.5 (51-60) |
| **max Age, years, median (IQR)** | 72 (69.75-79.25) |
| **Follow-up time, years, median (IQR)** | 20 (15-20) |
| **Duration of diagnosed HIV infection at the first sample, years, median (IQR)** | 9.3 (6.3-13.7) |
| **HIV viral load, median (IQR)** | 0 (0-0) |
| **Nadir CD4, median (IQR)** | 105.5 (39.75-195.75) |
| **nadir CD4 < 200, N (%)** | 41 (78.8%) |
| **Males, N (%)** | 44 (84.6%) |
| **White, N (%)** | 47 (90.4%) |
| **Ever smoked, N (%)** | 24 (46.2%) |
| **IDU, N (%)** | 3 (5.8%) |

Most individuals (N=38, 73%) had a single CHIP variant, 19% (N=10) had two variants and 8% (N=4) had three or four variants (Fig. 1C). The distribution of CHIP variants by gene followed the expected pattern (Figure 1D) with *DNMT3A* (N=28, 39%) being the most common followed by *TET2* (N=20, 28%) and *ASXL1* (N=12, 17%). The majority of CHIP variants were missense (N=26, 36%), nonsense (N=17, 24%), or frameshift deletions (N=17, 24%; Fig. 1D). Half of the trajectories (N=36, 50%) originated during the follow-up period.

### Analysis of CHIP longitudinal dynamics in PWH

To analyse the CHIP trajectories’ dynamics, for each trajectory, we fit a cubic smoothing spline of VAF vs. age; then, for each fitted trajectory, we find first and second derivatives to estimate longitudinal speed and acceleration, respectively (Methods, Figure 1b).

We observed that most CHIP trajectories were growing, i.e., that they had a mean VAF speed > 0 (Figure 2b). Gene-specific VAF speed in PWH follows the well-documented pattern in the literature, with *TET2*, *ASXL1*, and *PPM1D* showing faster expansion and *DNMT3A* and *TP53* showing slower expansion (p=0.007, Mann-Whitney U test), and mean VAF speed for each gene positively correlated with previously estimated gene-specific fitness(Fabre et al.,^11^ (one-sided Pearson’s correlation coefficient = 0.82, p=0.045)). Acceleration was more homogeneous, except for *TET2* variants, which had accelerating, while the other genes had decelerating VAF trajectories (Figure 2a-c).

**Figure 2.**
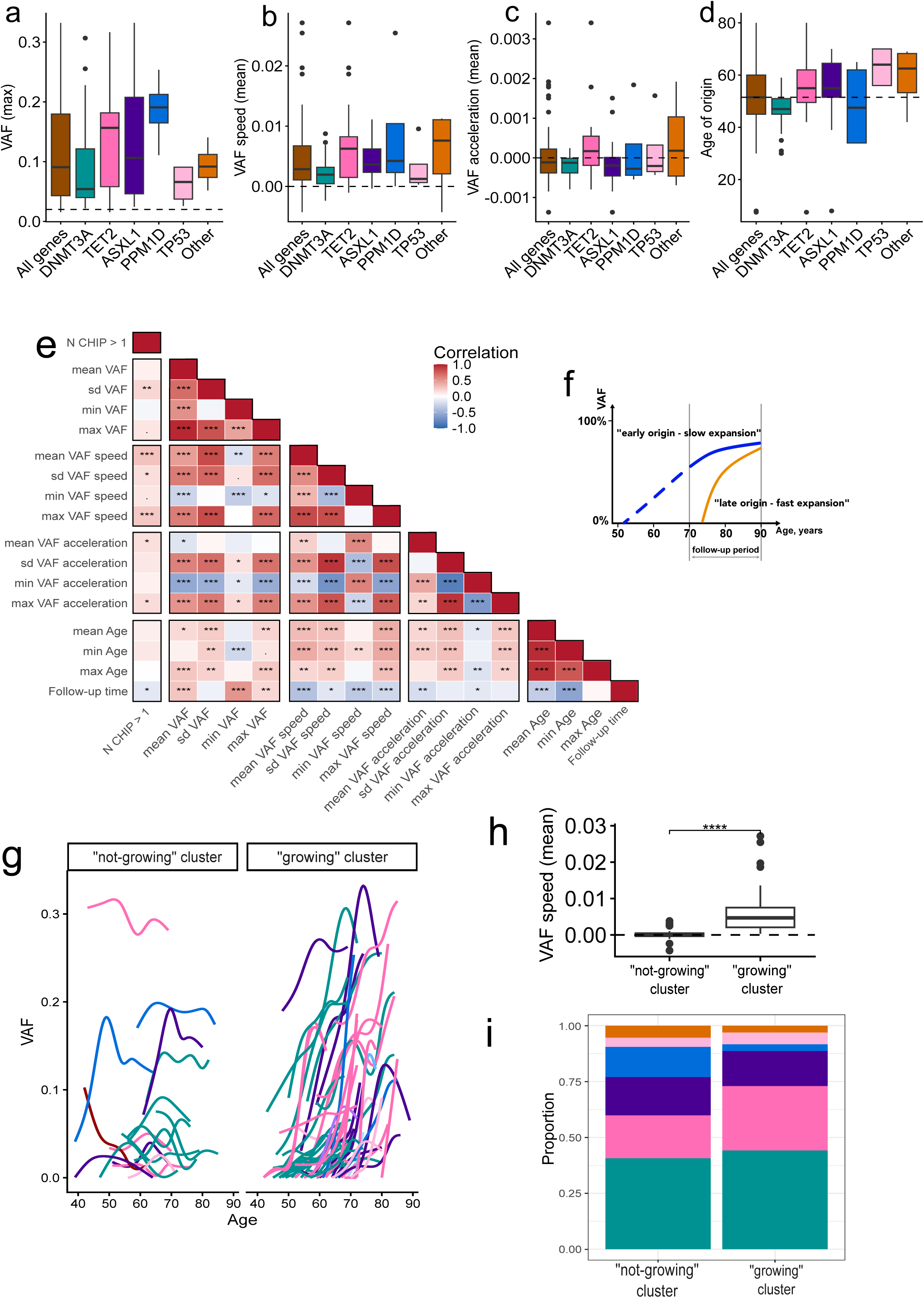
CHIP dynamics in PWH. a-d. Max VAF (a), mean VAF speed (b), mean VAF acceleration (c), and estimated age of origin (d) by CHIP gene. e. Pairwise correlation heatmap of VAF, VAF speed, VAF acceleration, and age summary statistics; speed/acceleration decrease as VAF increases, but are higher in late-onset trajectories. Color represents Spearman’s correlation coefficient, and starts represent the significance level: . - p<0.1, * - p<0.05, ** - p<0.01, *** - p<0.001. f. Scheme of CHIP clone dynamics deducted from the correlations in e. g. Clusters of VAF trajectories: not growing (left) and growing (right). h. Comparison of mean VAF speed of trajectories between trajectory clusters. i. Genes contributing to trajectory clusters (g). Colors in g and h correspond to genes and match colors in a-d.

We determined the age of origin of the trajectories using either the absence of the variant in a sequenced sample - for 36 trajectories with an origin during the follow-up period - or by linear extrapolation of the fitted splines, when possible - for the other trajectories (Figure 1b). The median age of origin was 51.5 years, with notable gene variation: variants in *DNMT3A* and *PPM1D* appeared at an earlier time point than those in *TP53* (Figure 2d).

To characterize the temporal dynamics of CHIP trajectories, we performed a systematic correlation analysis of key summary statistics (mean, min, max, SD; see Figure 2e) of VAF, VAF speed, VAF acceleration, and age across all trajectories. This analysis revealed that speed and acceleration tend to decrease, while VAF is positively associated with trajectory duration and subject age at observation. Paradoxically, trajectories recorded at more advanced ages demonstrated higher speed and acceleration. These divergent patterns suggest two distinct modes of clonal evolution: an “early-origin, slow-expansion” phenotype, characterized by clones that arise earlier in life and undergo gradual, protracted growth; and a “late-origin, fast-expansion” phenotype, characterized by clones that emerge at later ages and undergo rapid, accelerated proliferation (Figure 2f).

Multiple CHIP variants were observed in several individuals, allowing qualitative observation of co-expanding or competing clones. Co-occurring trajectories can represent subclones – i.e., two variants appearing in the same cells showing parallel trajectories – or competing clones – i.e., variants present in different cells, more likely to have divergent trajectories. Despite the fact that we lacked the power to perform a statistical analysis comparing co-occurring and competing clones, we can describe some patterns of those trajectories’ dynamics. For example, individuals SHCS_3, SHCS_32, and SHCS_42 each have two CHIP trajectories that appear to be in one clone (Figure 3 and Figure S1). For individuals with distinct clones, we see that one of the clones is outgrowing the other, such as in participant SHCS_37 (Figure 3), the first clone with a variant in *DNMT3A* decreased in size after the second variant in *JAK2* appeared and started to grow. The latter was also eventually outgrown by the third clone, which had 2 variants in *TET2*. These observations illustrate the complexity of clonal architecture in aging hematopoiesis.

**Figure 3.**
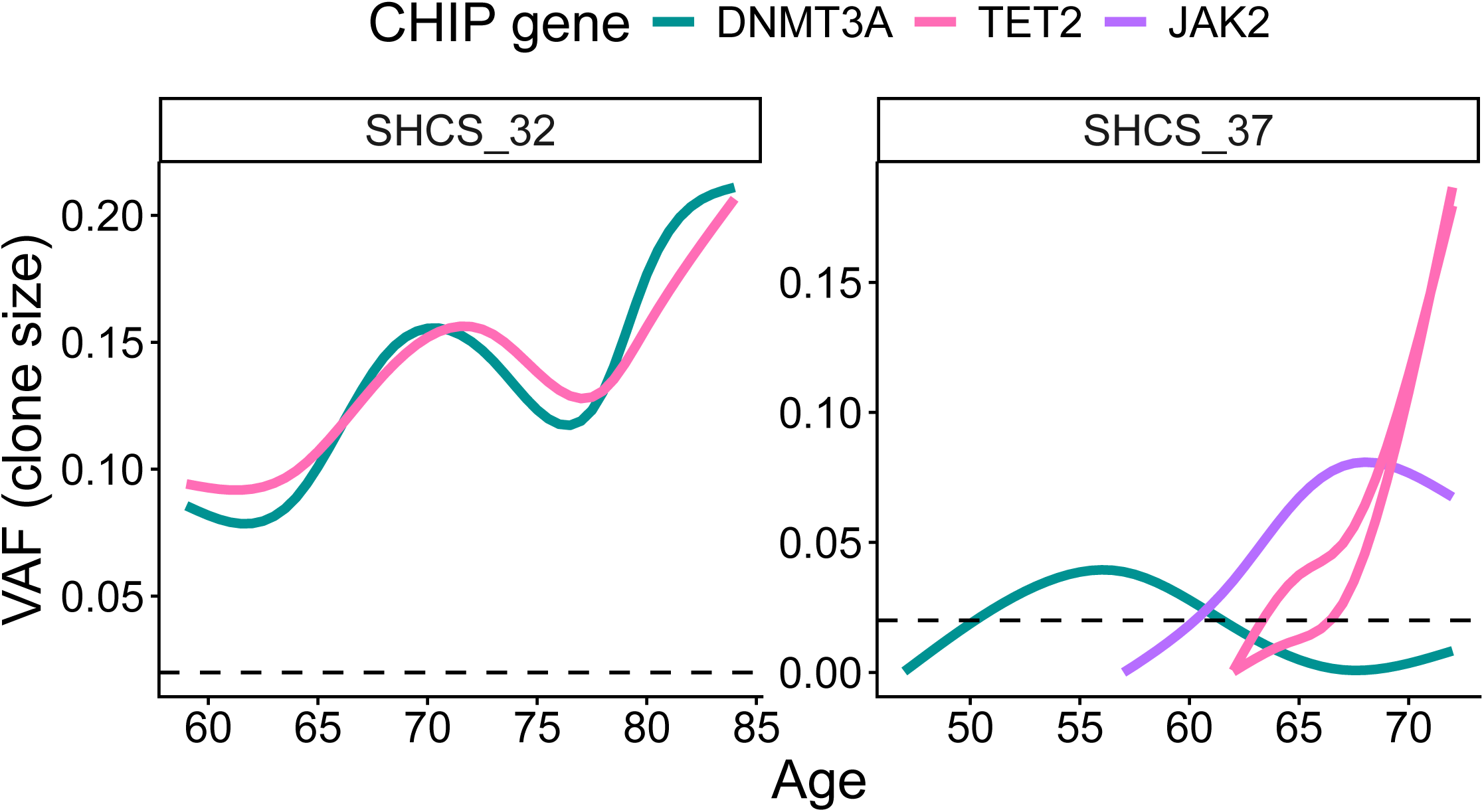
Examples of different types of co-occurring CHIP trajectories – subclones (participant SHCS_32), where two CHIP variants are present in the same cells and have matching longitudinal dynamics, and competing clones (participant SHCS_37), where distinct clones harbour distinct CHIP variants and show competing dynamics.

To examine trajectories in an unsupervised manner, we clustered them using dynamic time warping (DTW) distance and dynamic barycenter averaging (DBA) to define cluster centroids (see Methods). This analysis identified two main trajectory clusters, which we labeled “not-growing” (cluster 1, n = 17) and “growing” (cluster 2, n = 55; Figure 2g). The two clusters were similar in mean VAF, mean age, and acceleration, but trajectories in the “growing” cluster had higher VAF speed (Figure 2h) and an older age of origin (Figure S2), and a larger fraction of them originated during the follow-up period: 60% (N=33), vs. 18% (N=3) in the “not-growing” cluster. CHIP genes were distributed similarly across both clusters (Figure 2i). In addition, nearly all co-occurring trajectories in the “not-growing” cluster had a paired trajectory in the “growing” cluster (Figure S3). Several co-occurring trajectories in the “growing” cluster appeared to represent subclones, whereas no subclonal trajectories were observed in the “not-growing” cluster.

### Determinants of clonal growth in PWH

We sought to determine which demographic and HIV-related factors might affect longitudinal CHIP dynamics. We searched for associations between trajectory characteristics (VAF, speed, and acceleration, as in Figure 1) and the following factors (adjusted by age): sex, smoking status, injection drug use (IDU), nadir CD4 T-cell count, duration of untreated infection, and exposure to different ART regimens. We found that IDU was associated with larger clone size (p=0.02), and that longer period with detectable viral RNA was associated with higher acceleration (p=0.02; Table S1).

We also compared the two trajectory clusters with respect to demographic and HIV-related factors (Table S2). Individuals with trajectories in the “not-growing” cluster had more severe historical immune suppression compared with those with trajectories in the “growing” cluster. Specifically, the “not-growing” cluster included a higher proportion of individuals with a history of injection drug use (18% vs 4%, p = 0.08), lower median nadir CD4 count (41 vs 140 cells/µL, p = 0.07), and a greater proportion of individuals with profound immunodeficiency (nadir CD4 < 50 cells/µL: 53% vs 22%, p = 0.03). In addition, individuals in the “not-growing” cluster experienced a longer duration of diagnosed but not treated HIV infection (median 1,035 vs 184 days, p = 0.09). Although several associations did not reach formal statistical significance, the overall pattern suggests that individuals with trajectories in the “not-growing” cluster experienced more prolonged and severe immune suppression before ART initiation.

We then explored the longitudinal relationship between CHIP trajectories and immune parameters, including CD4 and CD8 T-cell count, and CD4:CD8 ratio - the markers of immune system status in PWH. We fitted linear mixed-effects models with subject-specific random intercepts and slopes to assess longitudinal associations between CHIP VAF, VAF speed, and VAF acceleration and the immune parameters, adjusting for age and the interaction between age and each immune parameter. Results are summarized in Figure 4 and Table S2. Larger clone size (higher VAF) was associated with lower CD4 T-cell counts and lower CD4:CD8 ratios, consistent with an association between impaired immune reconstitution and increased clonal expansion. Higher VAF was also associated with lower total leukocyte and platelet counts, suggesting that broader hematopoietic and inflammatory states may influence clonal burden. The strongest predictors of clonal growth dynamics (speed and acceleration) were T-cell composition measures, particularly the proportion of CD3+ lymphocytes among total lymphocytes, indicating that variation in immune cell distribution may be linked to changes in the rate of clonal expansion. Platelet count showed an opposite association with clonal kinetics, being negatively associated with VAF and speed but positively with acceleration. The association strength and direction mostly differed by gene (Figure S4) and by trajectory cluster (Figure S5), but the following associations were consistent across most genes and both clusters: lower CD4 count and CD4:CD8 ratio with higher clone size, higher proportion of CD3 cells with higher VAF speed, and lower platelet count with higher VAF speed.

**Figure 4.**
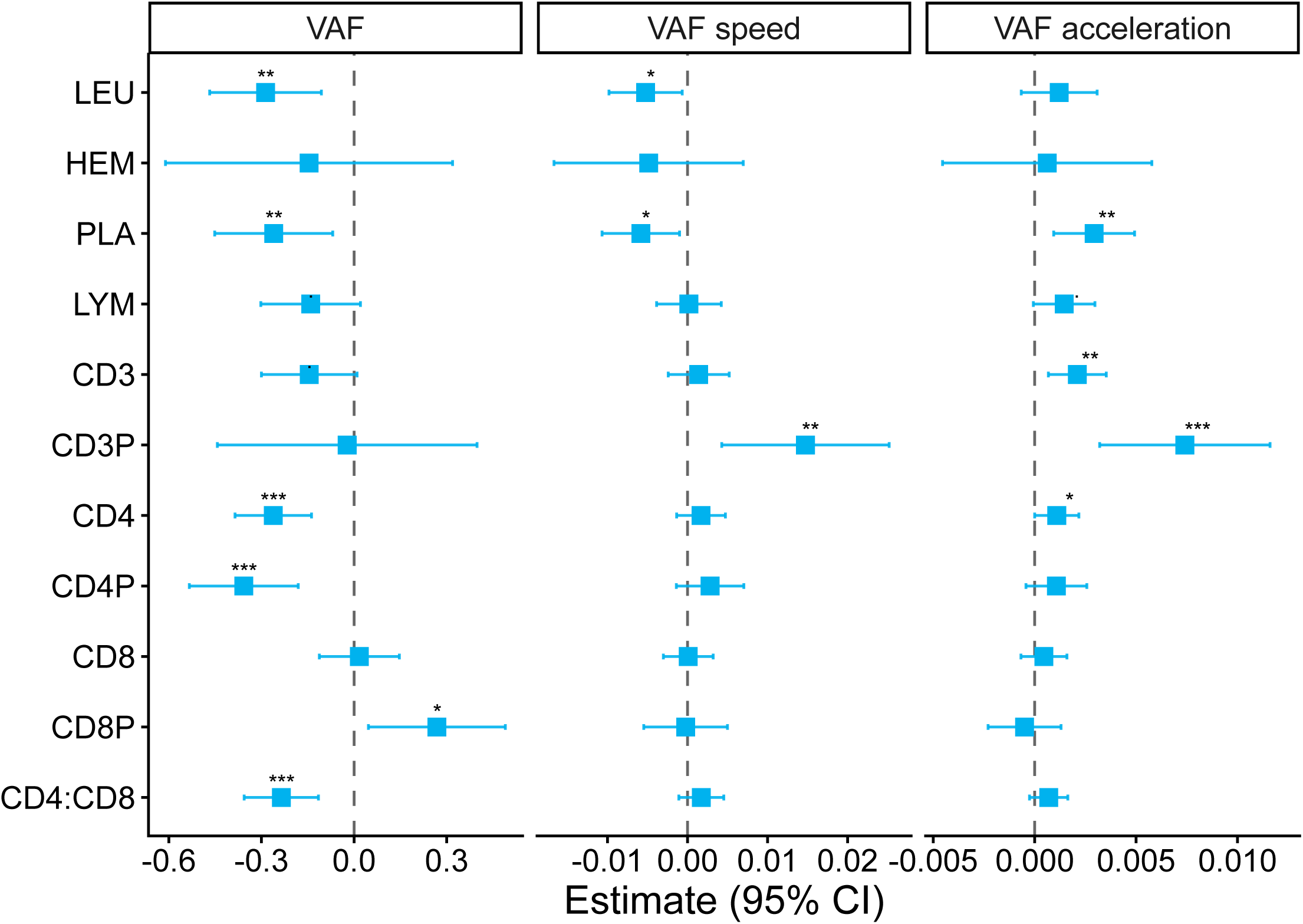
CHIP dynamics vs. blood parameters. Each coefficient represents a separate mixed linear model of the CHIP trajectory, with blood parameter as the predictor. Each model was adjusted for Age and the interaction term between Age and the blood parameter; only measurements taken after ART initiation were included. LEU - leukocyte count (cells/μl); HEM - hemoglobin (g/dl); PLA - platelet count (10^9^ cells/l); LYM - lymphocyte count (cells/μl) CD3 - CD3+ cell count (cells/μl); CD3P - CD3+ cells as a % of lymphocytes; CD4 - CD4+ cell count (cells/μl); CD4P - CD4+ cells as a % of lymphocytes; CD8 - CD8+ cell count (cells/μl); CD8P - CD8+ cells as a % of lymphocytes; CD4:CD8 - ration of CD4+ to CD8+ cell counts.

### Comparison with the general population cohort

For comparison between PWH and non-infected individuals, we compared SHCS participants (PWH) with publicly available CHIP longitudinal data from the SardiNIA cohort (general population)^11^. SardiNIA is a longitudinal study of individuals from four towns in Sardinia, with five sampling and data collection phases spanning more than 20 years. We filtered CHIP trajectories, fitted cubic splines, and derived VAF speed and acceleration in SardiNIA in the same manner as in SHCS. We restricted the analysis to the 5 most prevalent CHIP genes: *DNMT3A, TET2, ASXL1, PPM1D,* and *TP53.* Because age distribution during the observation period differed substantially between SHCS and SardiNIA, we applied several levels of trajectory filtering and matching to account for these differences in sensitivity analyses (see Methods and Figure S6).

First, we performed a mixed-model analysis in which VAF was modeled as a function of Age and HIV status, allowing for random intercepts and slopes for individual trajectories. Across different matching strategies, the intercept was consistently higher in PWH (Figure 5a, Figure S7), whereas the slope did not differ significantly (Figure S8). A higher intercept with a similar slope in PWH suggests that CHIP in PWH may originate earlier and reach a higher VAF at older ages, without necessarily exhibiting faster growth during the study observation period. These findings remained consistent in gene-specific models fitted separately for each of the five CHIP genes (Figure S9). After assigning SardiNIA trajectories to the SHCS-derived trajectory clusters (see Figure 2g), the higher intercept in PWH was observed in both clusters (Figure 5b,c), although the effect was not significant in the “not-growing” cluster and stronger in the “growing” cluster.

**Figure 5.**
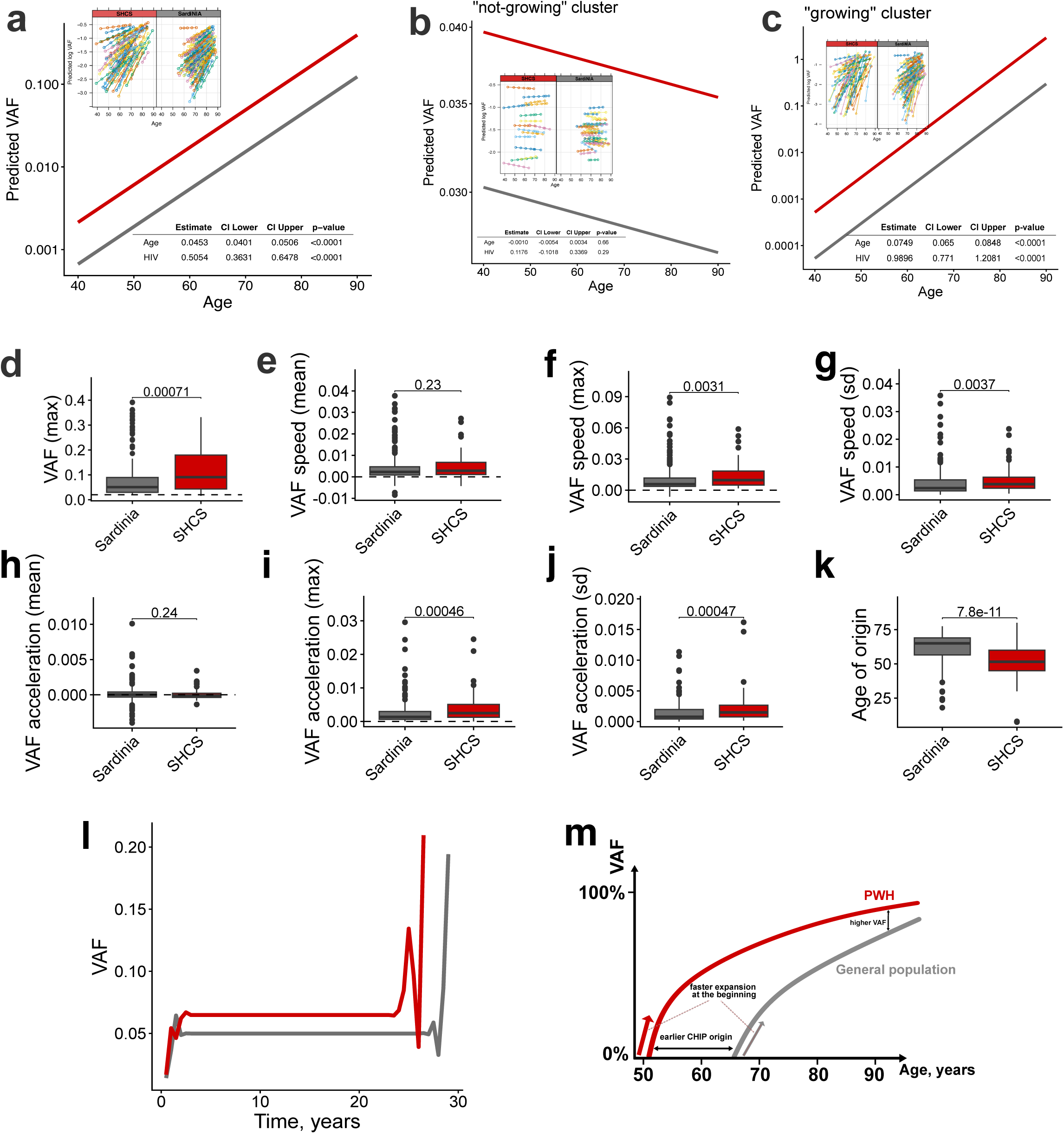
Comparison of CHIP trajectories between PWH (SHCS) and non-infected individuals (SardiNIA). a. The results and population-level prediction of a mixed linear model with random intercept and random slope for each CHIP trajectory of log(VAF) as a function of Age and HIV status in an unmatched set of trajectories (see Figure S5a). b-c. The results and population-level prediction of a cluster-specific mixed linear model with random intercept and random slope for each CHIP trajectory of log(VAF) as a function of Age and HIV status. d-k. The comparison of summary statistics of CHIP trajectories dynamics in an unmatched set of trajectories. l. Comparison of DBA barycenters of CHIP trajectories between SHCS and SardiNIA cohorts in an unmatched set of trajectories. m. The scheme of the proposed model of earlier origin and faster initial expansion of CHIP in PWH compared to the general population.

Consistent with the mixed model results, comparison of trajectory summary statistics showed no difference between PWH and the general population in mean VAF speed or acceleration. In contrast, PWH had higher maximal VAF speed, maximal VAF acceleration, maximal VAF, and an earlier inferred age of origin. Additionally, the SD of both speed and acceleration was greater in PWH, suggesting a stronger contribution of factors beyond genetically defined fitness of CHIP variants in this group (Figure 5d-k; Figure S10-13).

Finally, we applied DBA and visualised barycenters (consensus trajectories) for the two cohorts. This analysis showed a faster increase in VAF at the onset of the trajectory growth in PWH (Figure 5l), and the results were robust across matching strategies (Figure S14). Overall, these results support the model summarized in Figure 5m, in which CHIP appears earlier and expands more rapidly at the initial stage of trajectory growth in PWH.

## Discussion

In this study, we longitudinally reconstructed 72 clonal hematopoiesis trajectories in 52 PWH across more than two decades of follow-up and compared these dynamics with those observed in the general population. By combining spline-based trajectory reconstruction, dynamic time warping clustering, and mixed-effects modelling, we provide a detailed description of CHIP growth patterns and identify clinical and immunological factors associated with clonal expansion. Overall, our results show that CHIP dynamics in PWH are characterized by earlier emergence of detectable clones, faster expansion during the initial growth phase, and larger clone sizes later in life.

Consistent with previous longitudinal studies in the general population^11,12^, we observed substantial heterogeneity in clonal dynamics across CHIP genes. Somatic variants in *TET2*, *ASXL1*, and *PPM1D* generally showed faster expansion than variants in *DNMT3A* and *TP53*. This gene-specific pattern closely mirrors prior reports^11^ and supports the validity of our trajectory reconstruction approach.

Unsupervised clustering of trajectories identified two main dynamic patterns, corresponding broadly to “growing” and “non-growing” clones. Based on the comparison of dynamic parameters of two clusters (Figure S2), we suggest that trajectories in the “non-growing” cluster originated earlier in life and, by the observation period, either reached their maximal clone size or were outgrown by “growing” trajectories (Figure S3). These clusters were not strongly associated with specific driver genes, suggesting that trajectory shape reflects broader biological influences beyond mutation identity alone. Individuals with trajectories classified as “non-growing” tended to show evidence of more severe historical immune suppression, including lower nadir CD4 T-cell counts and longer duration of untreated HIV infection. Although these associations did not always reach statistical significance, their consistent direction suggests that immune suppression earlier in life, and especially during the untreated period of HIV-infection, may have lasting effects on clonal behaviour. In support of this interpretation, a larger CHIP clone size was associated with lower CD4 T-cell counts and lower CD4:CD8 ratios, both known markers of impaired immunity and chronic immune activation in PWH^27,28^. Measures of T-cell composition, particularly the proportion of CD3 lymphocytes, showed the strongest associations with clonal growth speed and acceleration, suggesting that immune cell distribution may influence clonal expansion dynamics. These findings are consistent with the hypothesis that inflammatory and immunosenescent environments confer a selective advantage to mutant hematopoietic stem cell clones^29,30^, and such phenotypes have been repeatedly associated with CHIP in PWH^20–22,31^. Alternatively, CHIP-associated mutations may themselves skew hematopoietic stem cell differentiation toward the myeloid lineage, thereby contributing to altered immune cell composition and reinforcing a feedback loop between clonal hematopoiesis and immune dysregulation^32–34^.

Comparison with the SardiNIA general population cohort^11^ provides important context for interpreting CHIP dynamics in PWH. Mixed-effects modelling showed that PWH had higher VAF values across the age range studied, while the rate of clonal expansion over that time was similar between cohorts. At the same time, maximal speed and acceleration were higher in PWH, and dynamic time warping barycenters showed a steeper increase in VAF early in trajectory development. The greater temporal variability in VAF speed and acceleration in PWH further suggests that individual and environmental factors may contribute more strongly to clonal fitness in this group than in the general population, where clonal growth is driven predominantly by mutation-specific genetic effects^11–13^. Together, these findings support a model in which chronic immune activation, inflammatory exposures, and other HIV-associated factors promote earlier clonal emergence or early expansion, while subsequent growth follows patterns broadly similar to those observed in the general population. This interpretation is consistent with prior evidence of accelerated biological ageing during untreated HIV infection, followed by partial normalization after initiation of effective antiretroviral therapy^35,36^.

This study has several limitations. First, the number of individuals with long-term serial samples was relatively small, reflecting the difficulty of obtaining longitudinal biospecimens spanning multiple decades. Second, participants in this study acquired HIV several decades ago and therefore experienced longer periods of untreated infection than contemporary patients, who typically initiate ART shortly after diagnosis. Third, although we sought to harmonize analyses between the SHCS and SardiNIA cohorts, differences in cohort structure, age distribution, and sequencing strategies may have introduced residual confounding.

Our findings contribute to understanding the mechanisms that may underlie the increased burden of age-related comorbidities observed in PWH. CHIP has been consistently associated with cardiovascular disease, chronic kidney disease, and other inflammatory conditions, supporting a role for mutant hematopoietic clones in promoting systemic inflammation and tissue dysfunction^37–39^. Persistent immune activation remains a hallmark of HIV infection, even in the era of effective ART, and has been implicated in premature aging and increased risk of non-AIDS morbidity^17,40^. The associations observed here between immune parameters and clonal dynamics suggest that immune dysregulation may represent an important mechanistic link between HIV infection and CHIP-associated morbidity.

Future studies combining longitudinal clinical data with multi-omics profiling will be important for clarifying how immune dysregulation shapes clonal evolution in PWH. In particular, single-cell genomic and transcriptomic approaches may help identify the specific immune cell populations harboring CHIP mutations and characterize their functional consequences, including altered inflammatory signaling, lineage bias, and immune effector programs^41–43^. Such approaches may help disentangle whether CHIP primarily acts as a driver of immune dysfunction, a consequence of chronic immune activation, or both. A better understanding of how HIV-associated immune perturbations interact with clonal hematopoiesis may ultimately improve risk stratification for age-related comorbidities in PWH and inform targeted strategies to mitigate inflammation-driven clonal expansion.

## Data Availability

The SHCS data are available to researchers upon the project submission and review by the Scientific Board of the SHCS and the study team; the provision of data is subject to Swiss legal and ethical regulations.

## Acknowledgements

This study has been financed within the framework of the Swiss HIV Cohort Study, supported by the Swiss National Science Foundation (grant #33FI-0_229621), by SHCS project #876, and by the SHCS research foundation. The data are gathered by the five Swiss University Hospitals, two Cantonal Hospitals, affiliated hospitals and private physicians.

## Members of the Swiss HIV Cohort Study

Abela IA, Aebi-Popp K, Anagnostopoulos A, Bernasconi E, Braun DL, Bucher HC, Calmy A, Cavassini M (Chairman of the Clinical and Laboratory Committee), Ciuffi A, Dollenmaier G, Egger M, Elzi L, Fehr JS, Fellay J, Frigerio Malossa S, Fux CA, Günthard HF, Hachfeld A, Haerry DHU (deputy of “Positive Council”), Hasse B, Hirsch HH, Hoffmann M, Hösli I, Huber M, Jackson-Perry D (patient representatives), Kahlert CR, Kaufmann D, Keiser O, Klimkait T, Kouyos RD, Kovari H, Kusejko K (Head of Data Centre), Labhardt ND, Leuzinger K, Martinez de Tejada B, Marzolini C, Metzner KJ, Müller N, Nemeth J, Nicca D, Notter J, Paioni P (Chairman of the Mother & Child Substudy), Pantaleo G, Perreau M, Rauch A (President of the SHCS), Salazar-Vizcaya LP, Schmid P, Segeral O, Speck RF, Stöckle M, Surial B, Tarr PE, Trkola A, Wandeler G (Chairman of the Scientific Board), Weisser M, Yerly S.

## Authorship Contributions

AGB performed targeted CHIP sequencing and CHIP variants annotation.

AC, MC, GC, HFG, LNW, PS, and PET contributed samples and data.

VT and JF performed the statistical analysis and interpreted the results.

All authors contributed to the study design and preparation of the manuscript.

