## Supplementary material for "Longitudinal modelling of clonal hematopoiesis reveals altered early clonal dynamics in people with HIV": Figure S

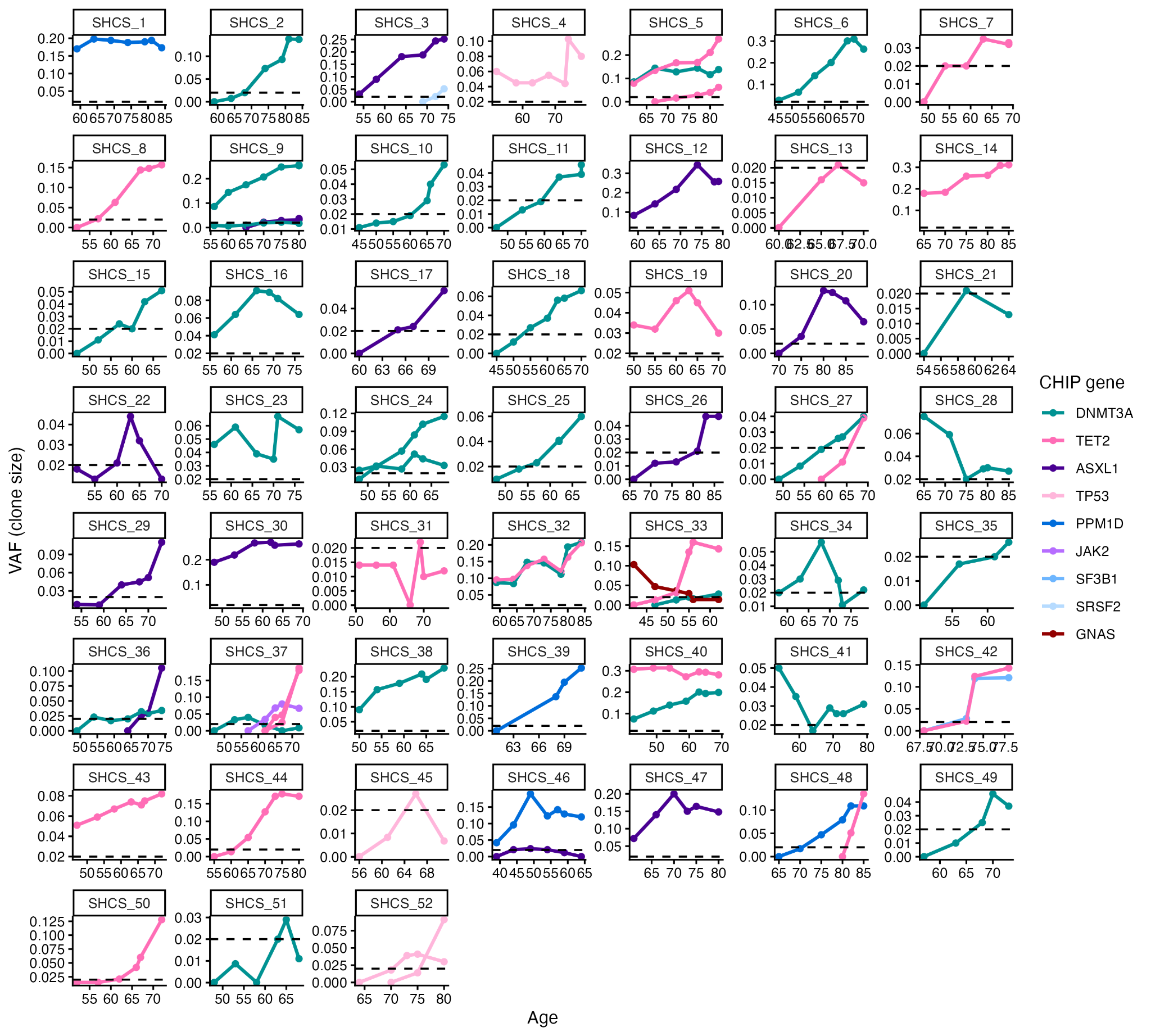

Figure S1. CHIP VAF trajectories analysed in the current study. Each panel represents one SHCS participant, and trajectories are colored according to the CHIP gene carrying the variant. The dashed line represents the VAF threshold of 2%.

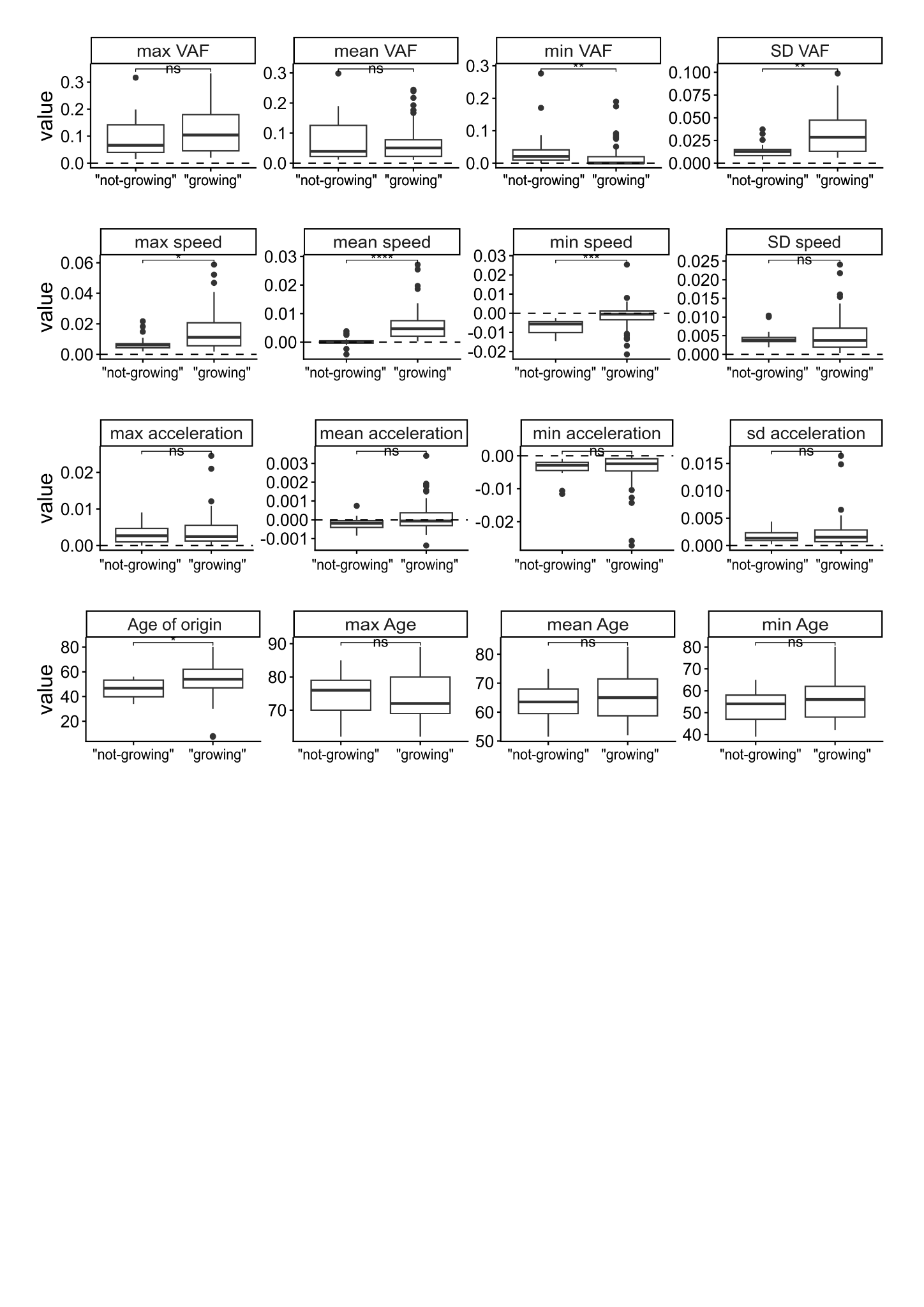

Figure S2. Comparison of trajectories’ summary statistics in two trajectory clusters (see Figure 2g).

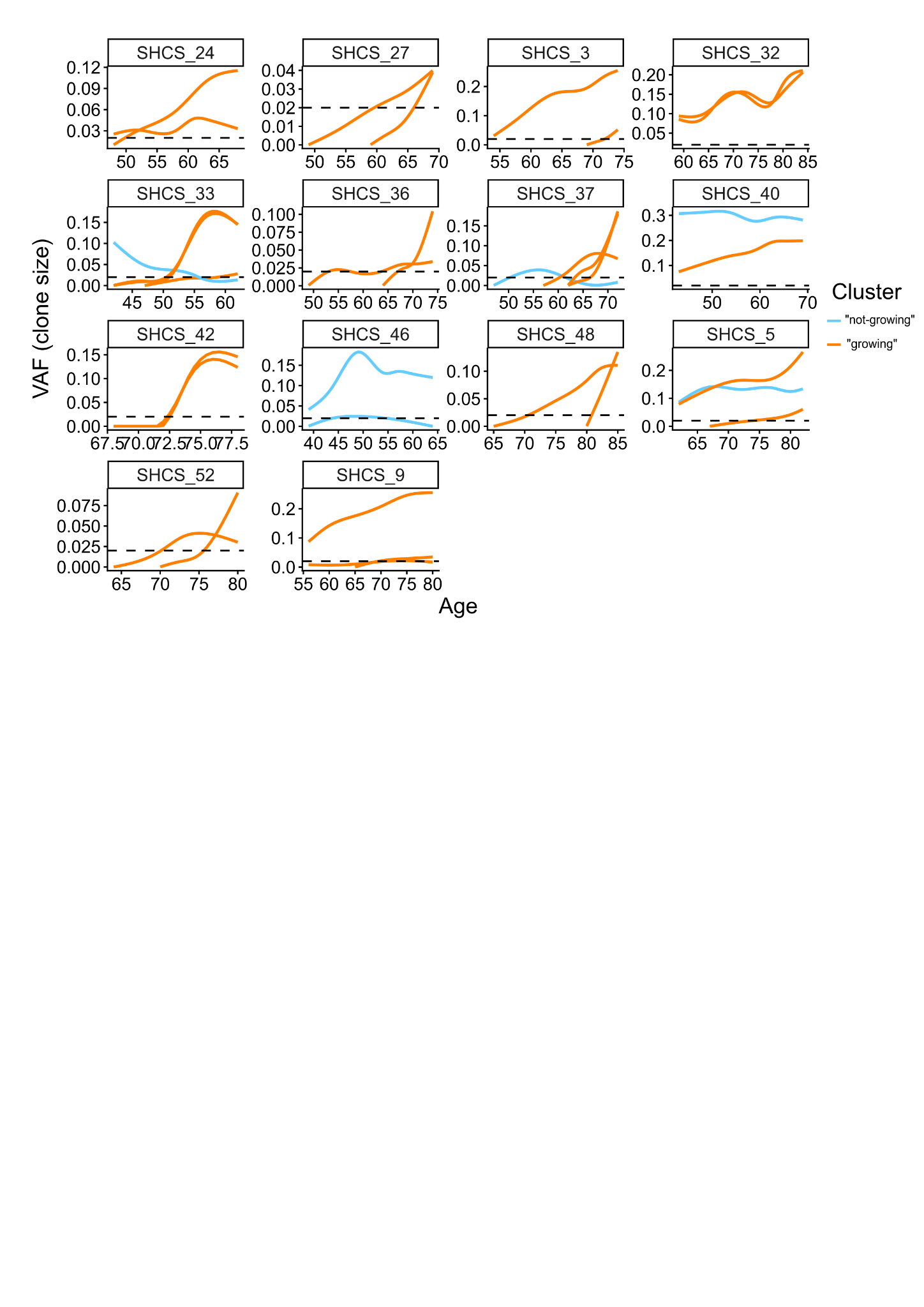

Figure S3. Individuals with several CHIP trajectories. Trajectories are colored according to the cluster (see Figure 2G).

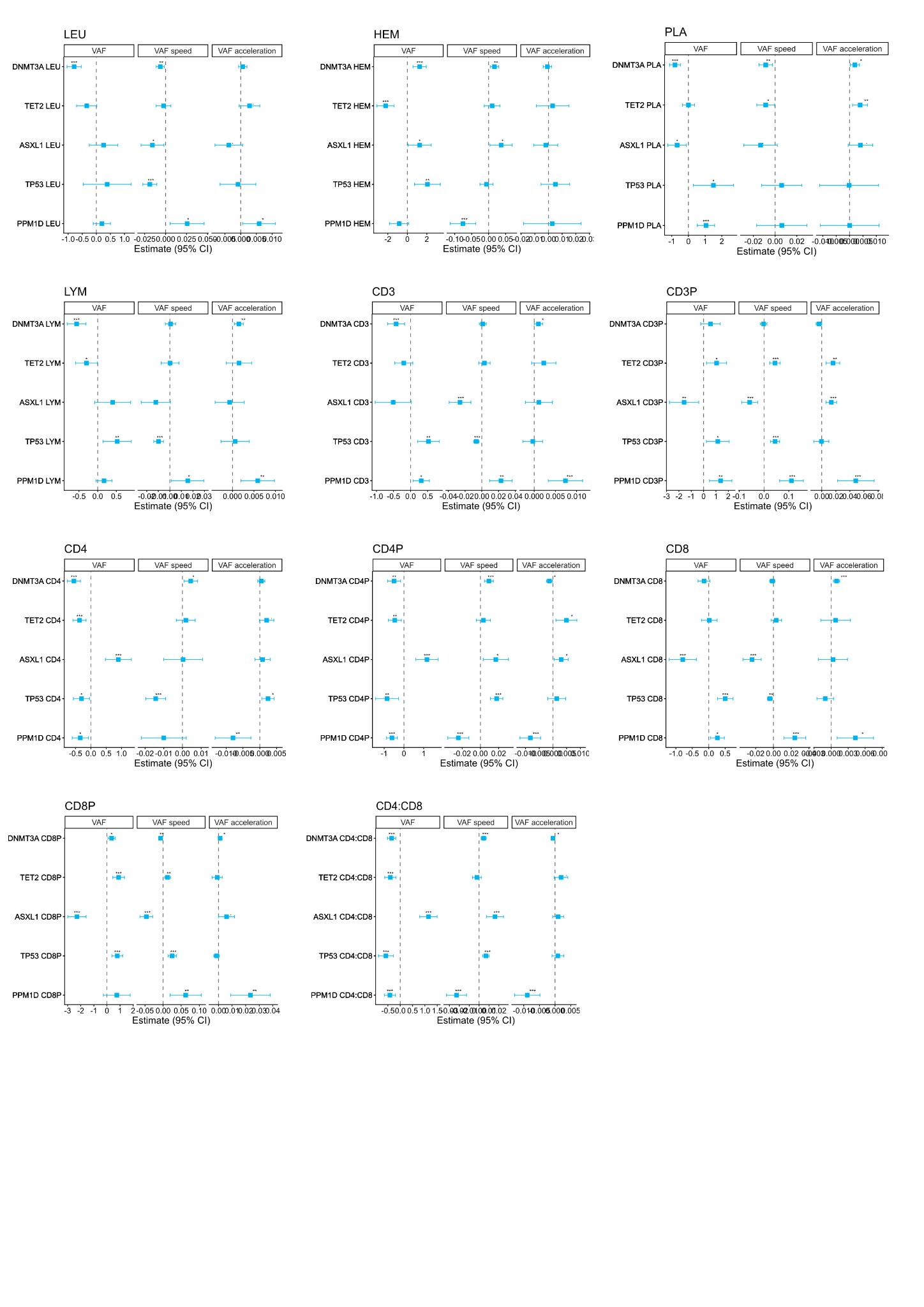

Figure S4. Gene-specific CHIP dynamics vs. blood parameters. Each coefficient represents a separate mixed linear model of the CHIP trajectory, with blood parameter as the predictor. Each model was adjusted for Age and the interaction term between Age and blood parameter; only measurements after ART start were included. LEU - leukocyte count (cells/μl); HEM - hemoglobin (g/dl); PLA - platelet count (10^9^ cells/l); LYM - lymphocyte count (cells/μl) CD3 - CD3+ cell count (cells/μl); CD3P - CD3+ cells as a % of lymphocytes; CD4 - CD4+ cell count (cells/μl); CD4P - CD4+ cells as a % of lymphocytes; CD8 - CD8+ cell count (cells/μl); CD8P - CD8+ cells as a % of lymphocytes; CD4:CD8 - ration of CD4+ to CD8+ cell counts.

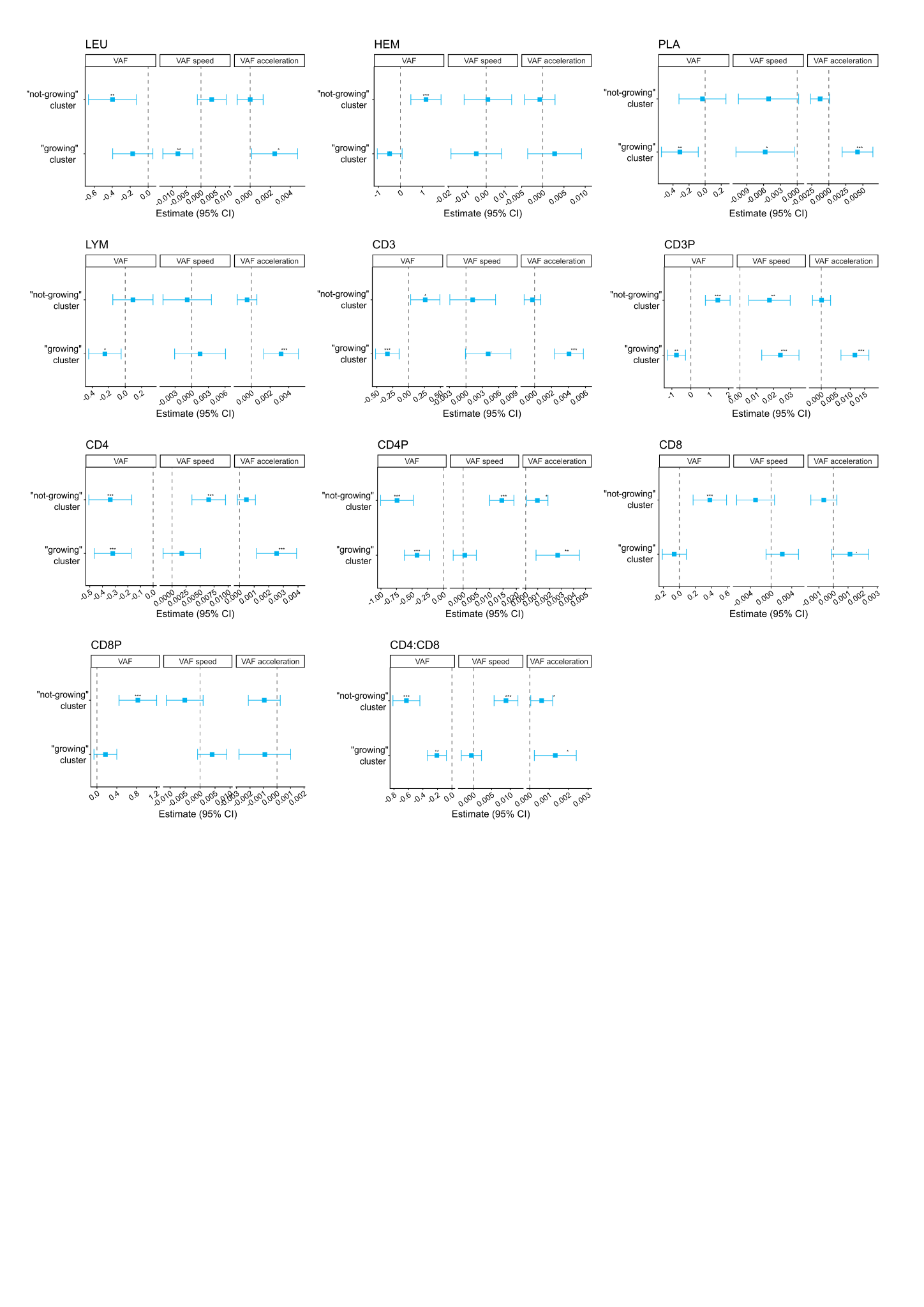

Figure S5. Cluster-specific CHIP dynamics vs. blood parameters. Each coefficient represents a separate mixed linear model of the CHIP trajectory, with blood parameter as the predictor. Each model was adjusted for Age and the interaction term between Age and blood parameter; only measurements after ART start were included. LEU - leukocyte count (cells/μl); HEM - hemoglobin (g/dl); PLA - platelet count (10^9^ cells/l); LYM - lymphocyte count (cells/μl) CD3 - CD3+ cell count (cells/μl); CD3P - CD3+ cells as a % of lymphocytes; CD4 - CD4+ cell count (cells/μl); CD4P - CD4+ cells as a % of lymphocytes; CD8 - CD8+ cell count (cells/μl); CD8P - CD8+ cells as a % of lymphocytes; CD4:CD8 - ration of CD4+ to CD8+ cell counts.

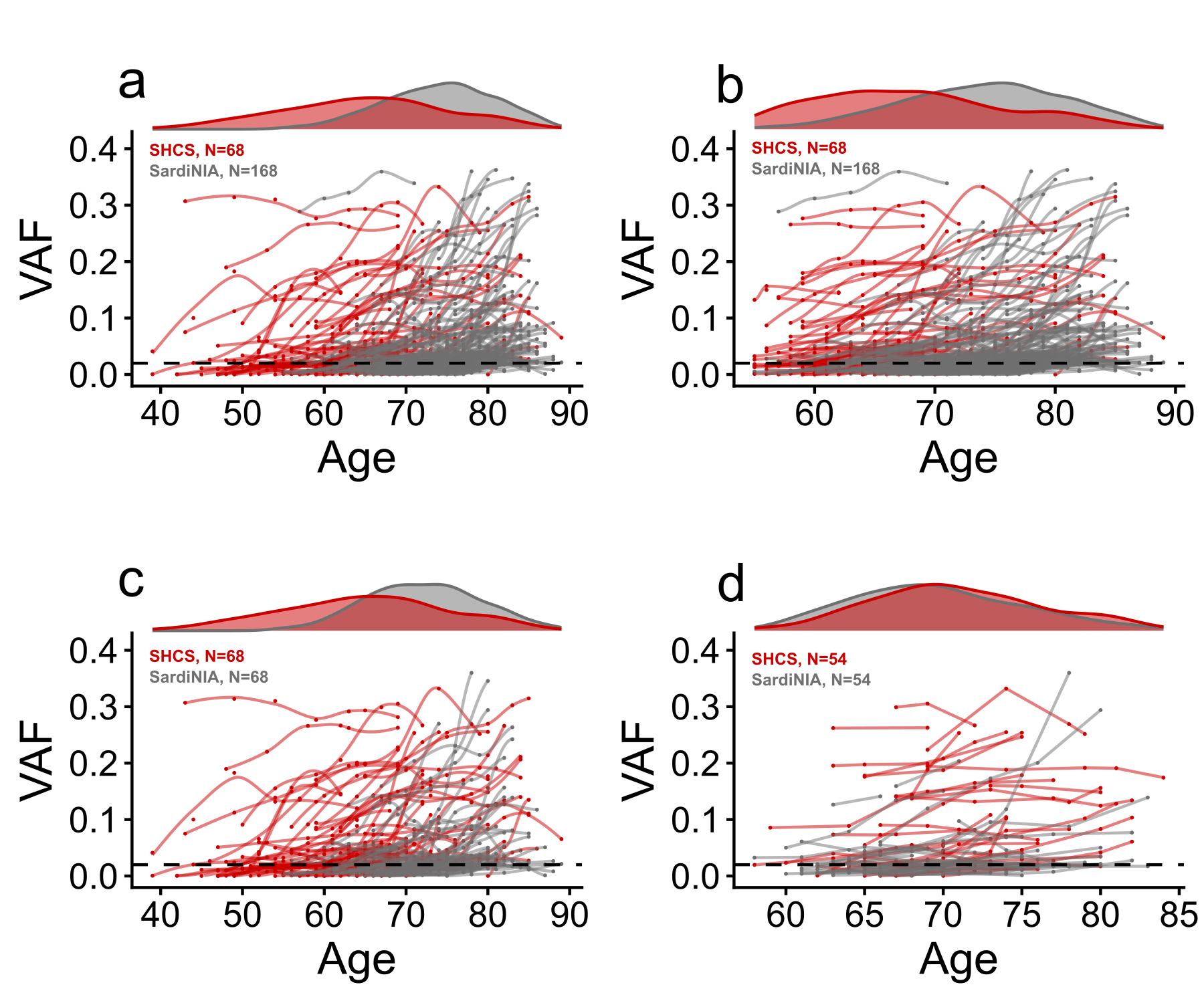

Figure S6. Different filtering and matching strategies of CHIP trajectories from SHCS and SardiNIA cohorts.

a. Unmatched and unfiltered - only common filters (at least 2 time points with a variant with one of them above 2% VAF, only common genes) are applied.

b. Unmatched and filtered - additionally, trajectories are filtered to keep time points at the age intersection between the two cohorts.

c. Matched and unfiltered - for each SHCS trajectory, we found the closest trajectory based on age in the same gene.

d. Matched and filtered - trajectories from panel c, filtered to keep time points at the age intersection between the two cohorts. Some trajectory pairs were excluded if the number of time points left was < 2.
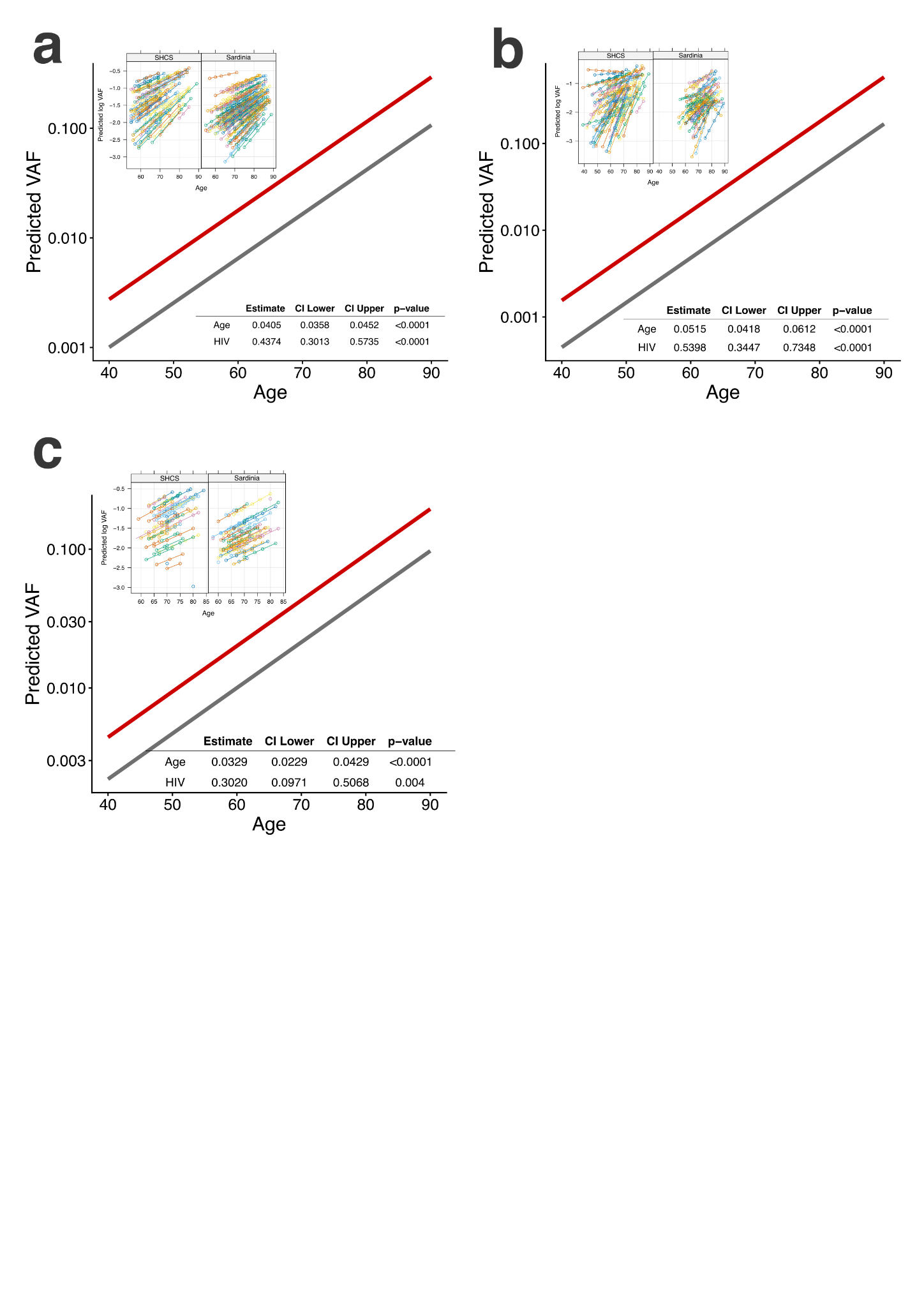

Figure S7. The results and prediction of the mixed linear model with random intercept and random slope of log(VAF) as a function of Age and HIV status in unmatched age-filtered (a), matched (b), and matched age-filtered (c) sets of trajectories. The y-axis is in log scale.

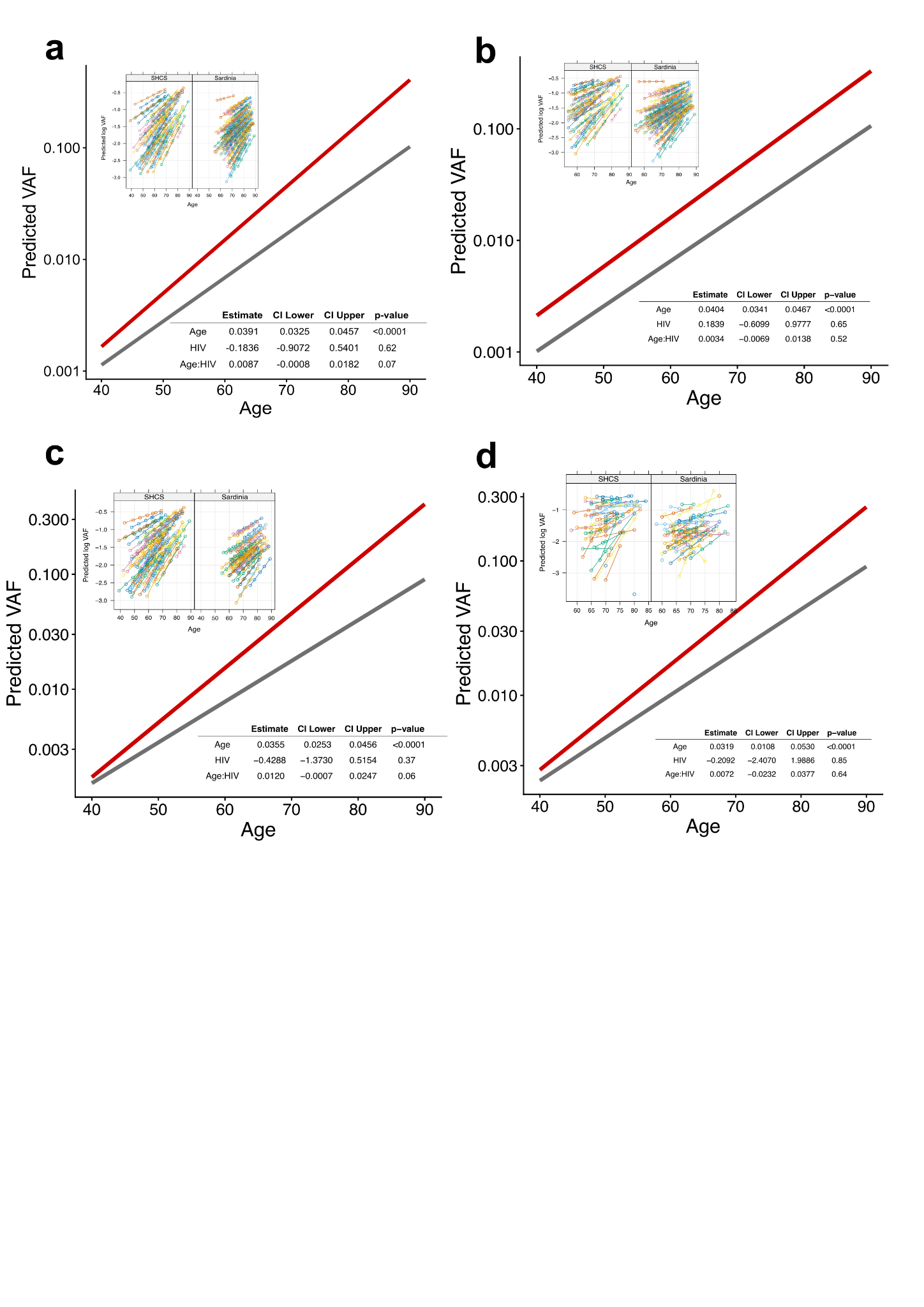

Figure S8. The results and prediction of the mixed linear model with random intercept and random slope of log(VAF) as a function of Age, HIV status, and their interaction in unmatched (a), unmatched age-filtered (b), matched (c), and matched age-filtered (d) sets of trajectories. The y-axis is in log scale.

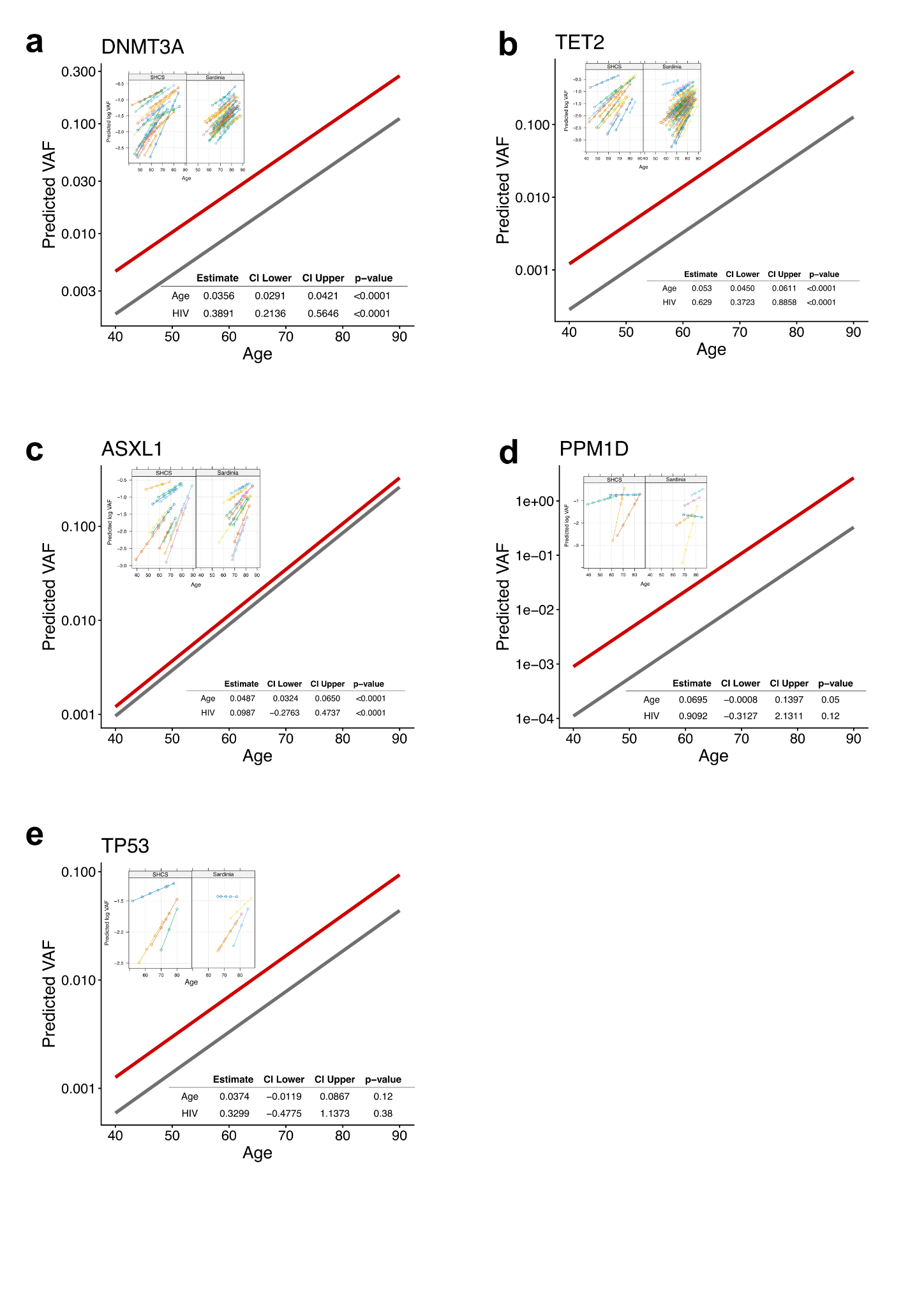

Figure S9. The results and prediction of the gene-specific mixed linear model with random intercept and random slope of log(VAF) as a function of Age and HIV status in the unmatched sets of trajectories. The y-axis is in log scale.

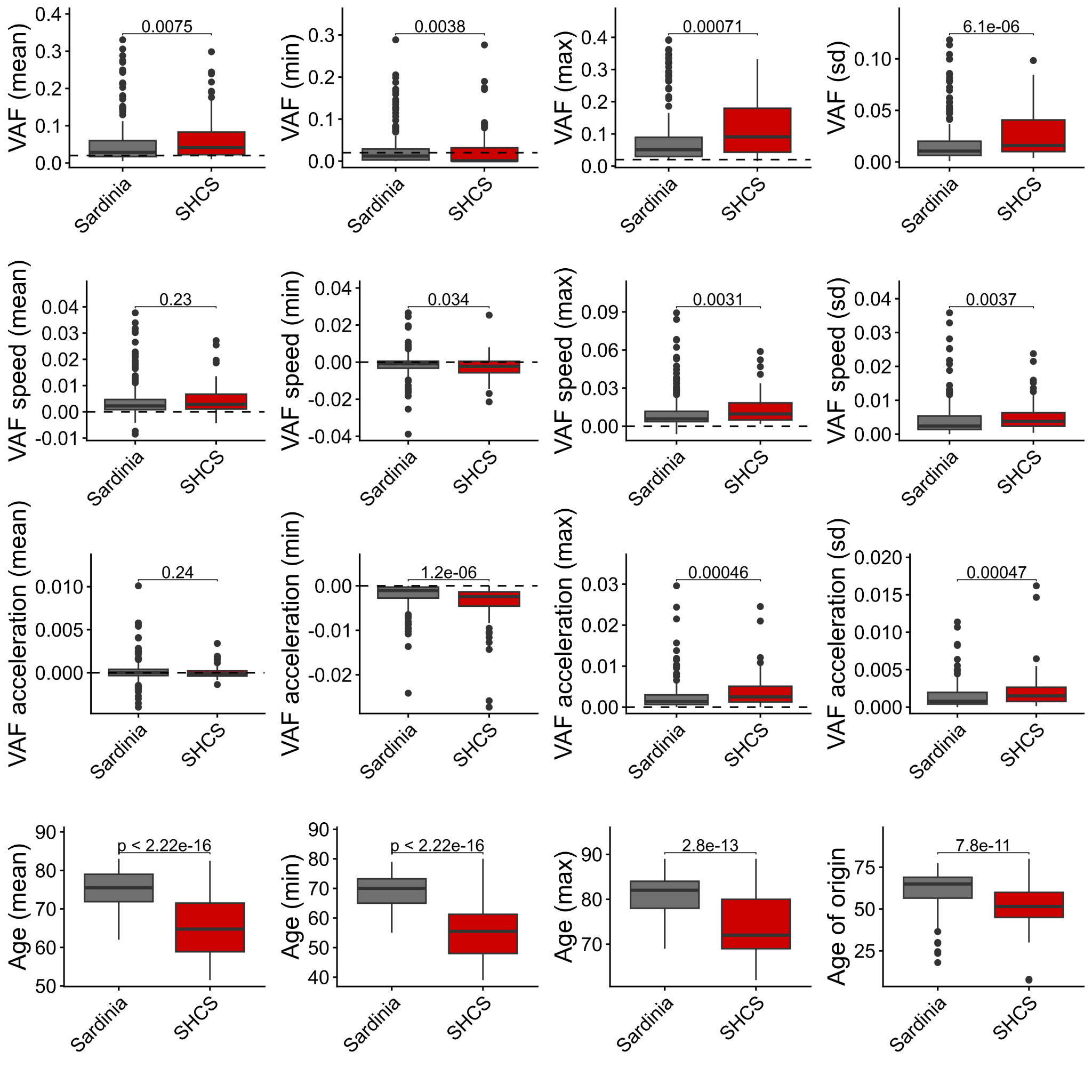

Figure S10. Comparison of CHIP trajectories’ summary statistics between SHCS and SardiNIA cohorts in an unmatched set of trajectories (Figure S5a).

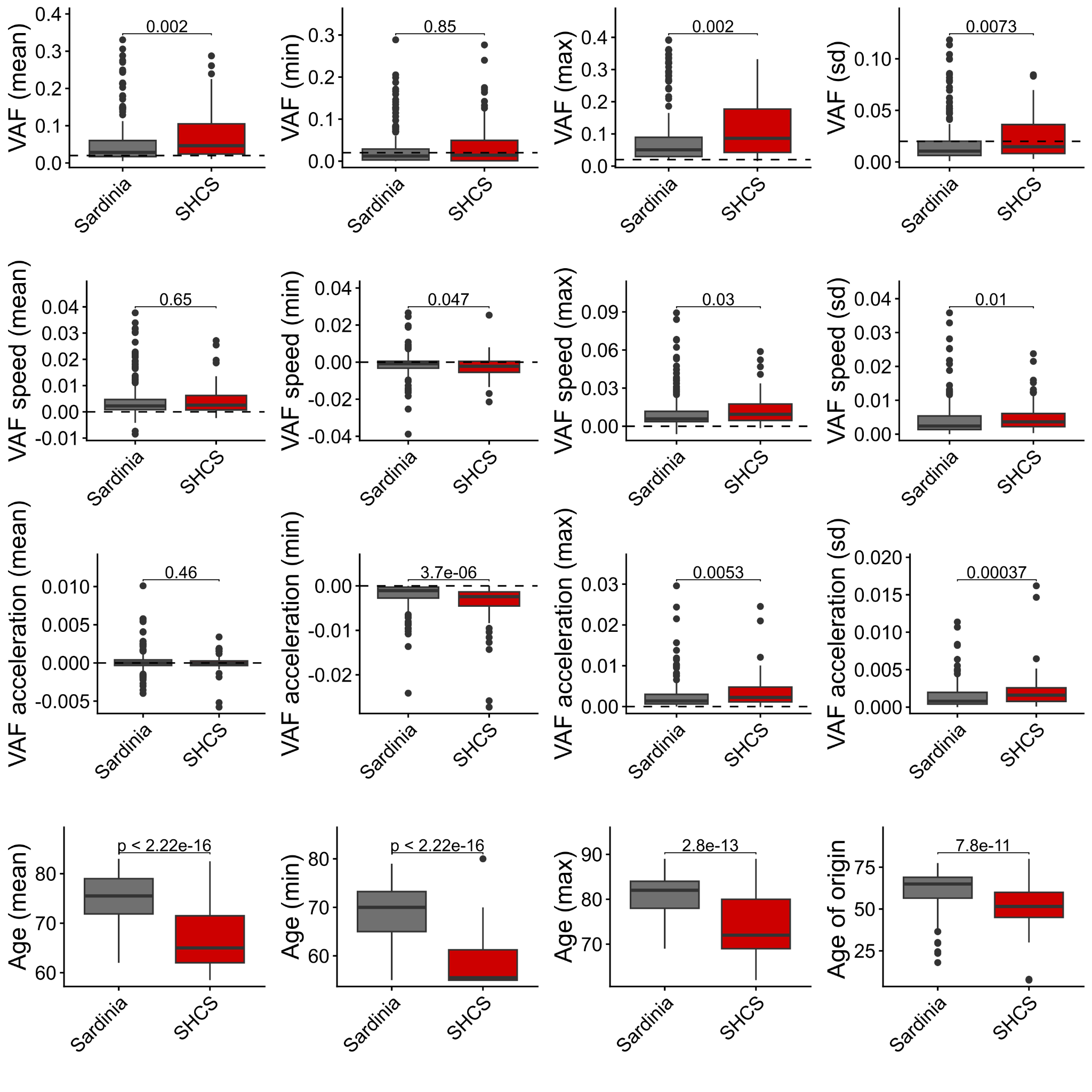
Figure S11. Comparison of CHIP trajectories’ summary statistics between SHCS and SardiNIA cohorts in an unmatched age-filtered set of trajectories (Figure S5b).

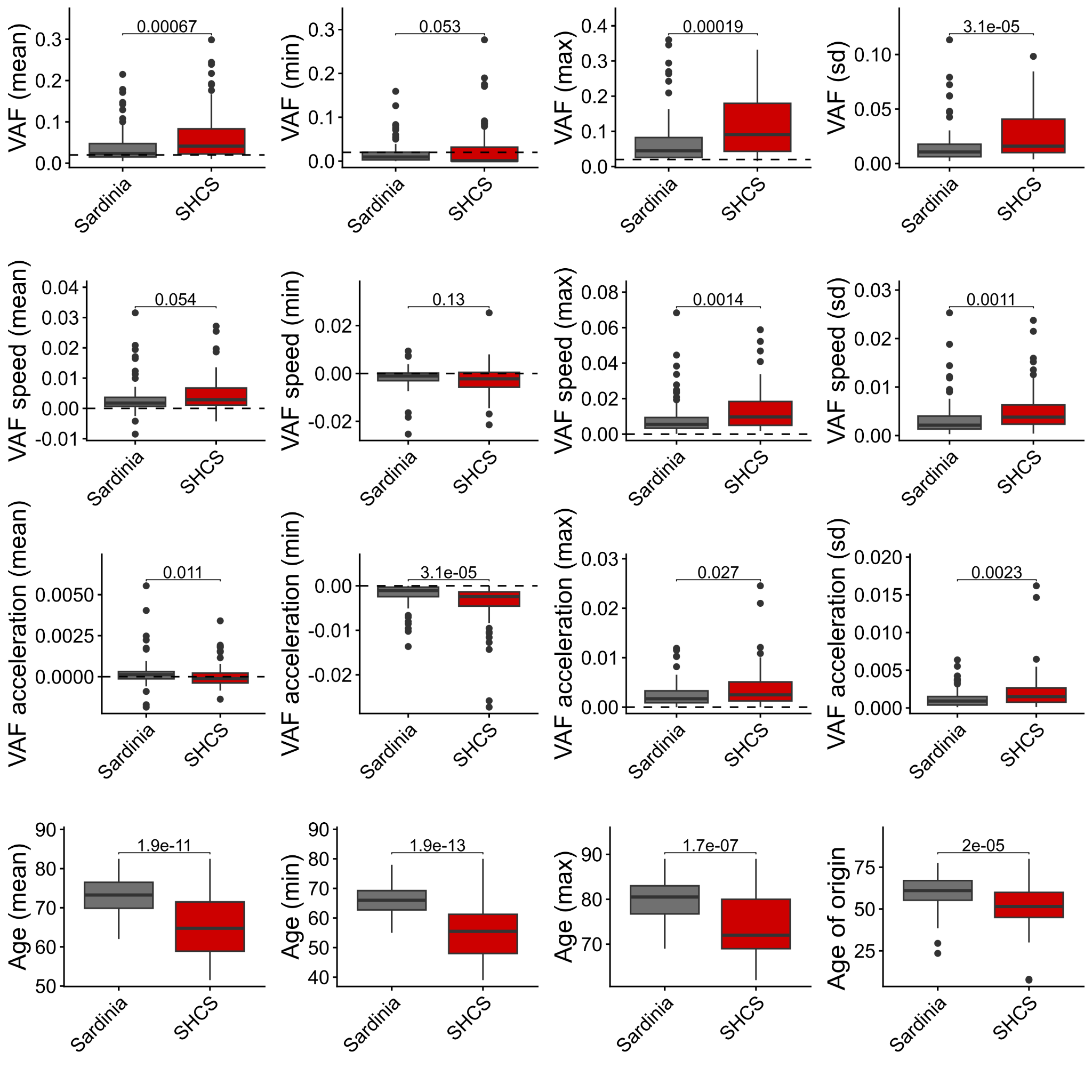

Figure S12. Comparison of CHIP trajectories’ summary statistics between SHCS and SardiNIA cohorts in a matched set of trajectories (Figure S5c).

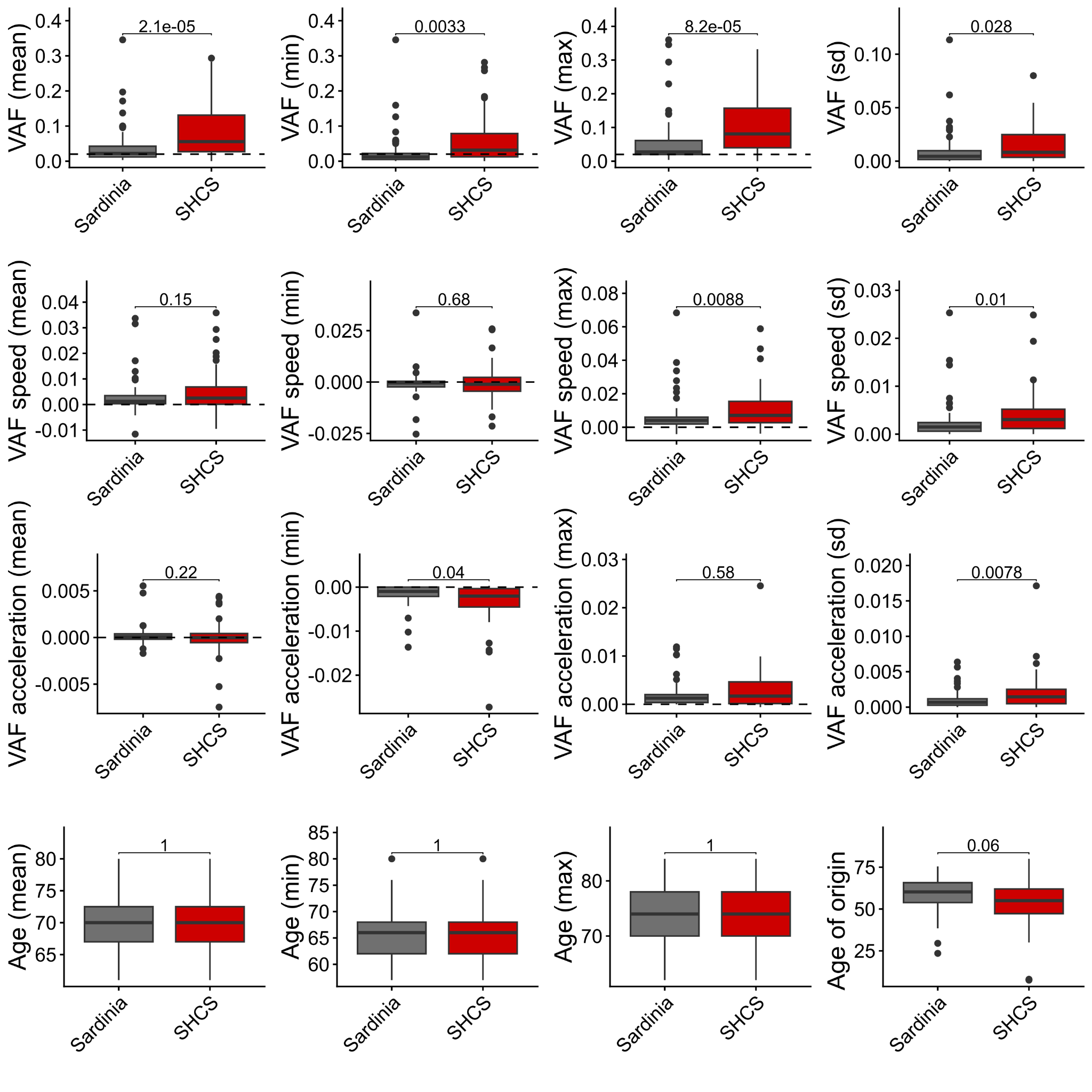

Figure S13. Comparison of CHIP trajectories’ summary statistics between SHCS and SardiNIA cohorts in a matched age-filtered set of trajectories (Figure S5d).

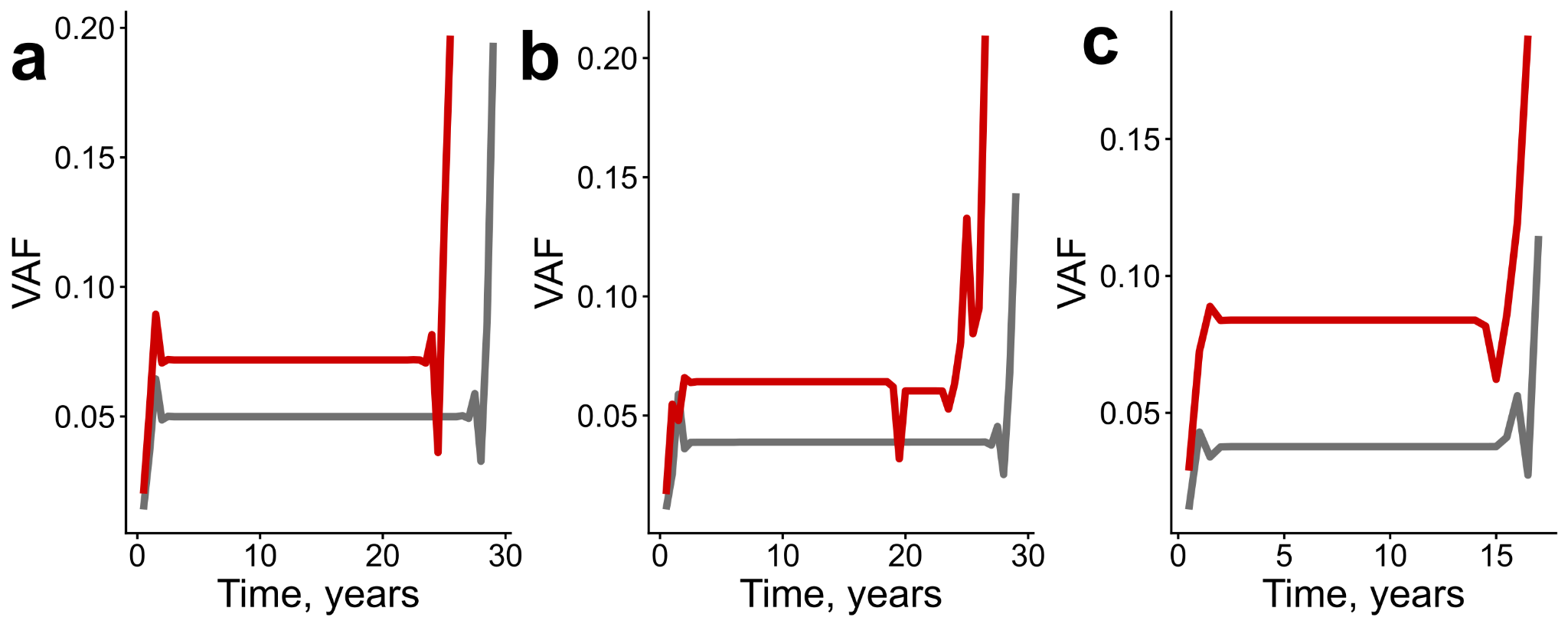

Figure S14. Comparison of DBA barycenters of CHIP trajectories between SHCS and SardiNIA cohorts in unmatched age-filtered (a), matched (b), and matched age-filtered (c) sets of trajectories.
